# Prenatal vitamin intake in first month of pregnancy and DNA methylation in cord blood and placenta in two prospective cohorts

**DOI:** 10.1101/2022.03.04.22271903

**Authors:** John F. Dou, Lauren Y.M. Middleton, Yihui Zhu, Kelly S. Benke, Jason I. Feinberg, Lisa A. Croen, Irva Hertz-Picciotto, Craig J. Newschaffer, Janine M. LaSalle, Daniele Fallin, Rebecca J. Schmidt, Kelly M. Bakulski

**Affiliations:** Department of Epidemiology, School of Public Health, University of Michigan, Ann Arbor, MI, USA; Department of Public Health Sciences and the M.I.N.D. Institute, School of Medicine, University of California, Davis, CA, USA; Department of Mental Health, Bloomberg School of Public Health, Johns Hopkins University, Baltimore, MD, USA; Division of Research, Kaiser Permanente, Oakland, CA, USA; College of Health and Human Development, Penn State University, State College, PA, USA

**Keywords:** DNA methylation, prenatal vitamins, epigenetics, epidemiology, pregnancy cohort, cord blood, placenta

## Abstract

**Background:** Prenatal vitamin use is recommended before and during pregnancies for normal fetal development. Prenatal vitamins do not have a standard formulation, but many contain calcium, folic acid, iodine, iron, omega-3 fatty acids, zinc, and vitamins A, B6, B12, and D, and usually they contain higher concentrations of folic acid and iron than regular multivitamins in the U.S. Nutrient levels can impact epigenetic factors such as DNA methylation, but relationships between maternal prenatal vitamin use and DNA methylation have been relatively understudied. We examined use of prenatal vitamins in the first month of pregnancy in relation to cord blood and placenta DNA methylation in two prospective pregnancy cohorts: the Early Autism Risk Longitudinal Investigation (EARLI) and Markers of Autism Risk Learning Early Signs (MARBLES) studies.

**Results:** In placenta, prenatal vitamin intake was marginally associated with -0.52% (95% CI: - 1.04, 0.01) lower mean array-wide DNA methylation in EARLI, and associated with -0.60% (−1.08, -0.13) lower mean array-wide DNA methylation in MARBLES. There was little consistency in the associations between prenatal vitamin intake and single DNA methylation site effect estimates across cohorts and tissues, with only a few overlapping sites with correlated effect estimates. However, the single DNA methylation sites with p-value<0.01 (EARLI cord n_CpGs_=4,068, EARLI placenta n_CpGs_ =3,647, MARBLES cord n_CpGs_ =4,068, MARBLES placenta n_CpGs_=9,563) were consistently enriched in neuronal developmental pathways.

**Conclusions:** Together, our findings suggest that prenatal vitamin intake in the first month of pregnancy may be related to lower placental global DNA methylation and related to DNA methylation in brain-related pathways in both placenta and cord blood.

## Introduction

Vitamins and minerals are critical for normal fetal development. The World Health Organization recommends supplementation during pregnancy with iron, folic acid, vitamin A, calcium, and iodine.(1) The American College of Obstetricians and Gynecologists additionally recommends supplementation with choline and vitamins B6, B12, C, and D.(2) Prenatal vitamins do not have a standard formulation, but most contain calcium, iodine, omega-3 fatty acids, zinc, and vitamins A and D as well as more iron and B vitamins, and about twice as much folic acid compared to multivitamins.(3–6) In the US, prenatal vitamin use among pregnant people is estimated to be between 78% and 92%, with 55-60% reporting use in the first trimester.(7–10) In another study of the EARLI cohort, 59.7% reported prenatal vitamin use in the first month.(6) Despite the use of these supplements, a recent study found that a significant number of pregnant people in the US still do not meet the recommended nutrient intake levels.(11) Deficiencies in these nutrients are associated with multiple disorders including anemia and preeclampsia in the parent and impaired neurodevelopment, neural tube defects, and recurrent wheezing in the child.(4,12–14) Understanding the molecular implications of these nutrients on fetal development is critical.

Nutrient levels can impact epigenetic factors such as DNA methylation. Relationships between prenatal vitamin use and DNA methylation have been relatively understudied, as most prior research focused on individual nutrients. For example, folic acid supplementation is associated with DNA methylation differences in cord blood at both differentially methylated regions(15) and differentially methylated positions in an epigenome-wide meta-analysis (n=1,988).(16) Another epigenome-wide association study examining vitamin D levels in two cohorts (n=1,416) found no association with cord blood DNA methylation.(17) Maternal dietary intake of three dietary patterns was not associated with DNA methylation in placental tissue, implying that minor nutritional deficiencies were not associated with differences in DNA methylation (n=573).(18) Conversely, a different study found that prenatal vitamin supplementation was associated with cord blood DNA methylation (n=130).(19) A systematic review of randomized control trials of maternal micronutrient supplementation and DNA methylation in cord blood, blood spots, placental tissue, and buccal swabs found inconsistent results, but these studies varied in exposure timing and dose, types of micronutrients, sample size, and analytical methods.(20) Additional studies of the associations between prenatal vitamin use and DNA methylation are needed.

Cord blood and placental tissues are useful sources of information to understand fetal development. Epigenetic factors such as DNA methylation play a role in normal and abnormal development.(21) DNA methylation patterns from cord blood and placental tissues may be used as biomarkers of *in utero* exposures or to predict future health.(22) DNA methylation has tissue-specific patterns, and tissues can have varying sensitivity to nutrient changes. Both cord blood and placental tissue develop early in gestation which represents an important window for early prenatal vitamin supplementation. The umbilical cord begins to form during the fourth week of gestation and blood is flowing in it by the fifth week, but the structure is not fully developed until the 12^th^ week.(23) Umbilical cord blood contains a higher level of hematopoietic stem cells compared to adult blood and includes differentiated cell types such as B cells, natural killer cells, T cells, monocytes, granulocytes, and nucleated red blood cells.(24,25) The placenta is the site of gas, nutrient, and waste exchange between the parent and fetus and has metabolic and endocrine functions. The trophoblastic cells of the placenta are of fetal origin, but these are in close contact with the maternal decidua and blood vessels through the chorionic villi.(26) The placenta starts to develop from the trophectoderm layer of the blastocyst following implantation in the maternal endometrium, around 6-7 days.(27)

The associations between prenatal vitamin use in the first month of pregnancy and DNA methylation in cord blood and placental tissue are still unclear. Here, we examined use of prenatal vitamin use during the first month of pregnancy and DNA methylation in cord blood and placenta in two prospective pregnancy cohorts. We analyzed array-wide mean DNA methylation, as well as associations at single DNA methylation sites and tested for enriched gene ontology processes.

## Results

### Study Sample Descriptive Statistics

For cord blood, variables of interest (prenatal vitamin use, maternal education, maternal age, genetic principal components, sex, gestational age, estimated cell proportions) were available for 113 samples (66.5% of total) in EARLI and 201 samples (82.7% of total) in MARBLES. For placenta, variables of interest were available for 88 samples (69.3% of total) in EARLI and 70 samples (77.8% of total) in MARBLES. Analyses were done separately in the four cohort/tissue groups: EARLI cord blood, EARLI placenta, MARBLES cord blood, and MARBLES placenta.

Among subjects with cord blood samples, use of prenatal vitamins in the first month of pregnancy was associated with maternal education in both cohorts. In EARLI, 69.0% of mothers who took prenatal vitamins in the first month had a college degree, compared to 42.9% of mothers who did not take prenatal vitamins. In MARBLES, 63.7% of mothers who took prenatal vitamins in the first month of pregnancy had at least a college degree, while 34.3% of mothers who did not take prenatal vitamins had a college degree (**Table 1**). Among the subset with placenta samples, this same trend was less pronounced in EARLI (64.2% mothers who took prenatal vitamins had college degree versus 51.4% among mothers who did not take prenatal vitamins), and present in MARBLES (69.7% taking prenatal vitamins in first month had college degree compared to 43.2% among those without prenatal vitamin use) (**Table 2**).

**Table 1.** Distributions of maternal and fetal participant and sample characteristics for those with cord blood DNA methylation measures, split by maternal use of prenatal vitamins in month 1 of pregnancy.

|  | MARBLES |  |  | EARLI |  |  |
| --- | --- | --- | --- | --- | --- | --- |
|  | No use of prenatal vitamins month 1 | Use of prenatal vitamins month 1 | P-value | No use of prenatal vitamins month 1 | Use of prenatal vitamins month 1 | P-value |
|  | N=99 | N=102 |  | N=42 | N=71 |  |
| **Maternal education** |  |  | <0.001 |  |  | 0.01 |
| **No degree** | 65 (65.7%) | 37 (36.3%) |  | 24 (57.1%) | 22 (31.0%) |  |
| **College degree** | 34 (34.3%) | 65 (63.7%) |  | 18 (42.9%) | 49 (69.0%) |  |
| **Maternal age at delivery (years)** | 34.8 (4.95) | 34.0 (4.67) | 0.264 | 33.7 (4.48) | 33.1 (4.46) | 0.51 |
| **Child race/ethnicity** |  |  | 0.626 |  |  | 1.00 |
| **Non-Hispanic White** | 41 (41.4%) | 44 (43.1%) |  | 22 (52.4%) | 36 (50.7%) |  |
| **Hispanic** | 35 (35.4%) | 30 (29.4%) |  | 5 (11.9%) | 9 (12.7%) |  |
| **Other/Multiple** | 23 (23.2%) | 28 (27.5%) |  | 15 (35.7%) | 26 (36.6%) |  |
| **Child sex** |  |  | 0.583 |  |  | 0.08 |
| **Female** | 37 (37.4%) | 43 (42.2%) |  | 25 (59.5%) | 29 (40.8%) |  |
| **Male** | 62 (62.6%) | 59 (57.8%) |  | 17 (40.5%) | 42 (59.2%) |  |
| **Gestational age at delivery (weeks)** | 39.2 (1.15) | 39.1 (1.27) | 0.519 | 39.1 (1.49) | 39.5 (1.33) | 0.24 |
| **Estimated cell proportion** |  |  |  |  |  |  |
| **CD8T** | 4.11 (2.89) | 3.66 (2.75) | 0.26 | 3.51 (2.55) | 3.99 (3.00) | 0.37 |
| **CD4T** | 17.4 (6.08) | 17.8 (5.66) | 0.66 | 17.6 (8.36) | 17.1 (7.31) | 0.74 |
| **Natural Killer** | 3.25 (2.90) | 3.05 (2.73) | 0.62 | 3.39 (2.48) | 3.71 (2.93) | 0.53 |
| **B Cell** | 5.62 (2.88) | 5.36 (2.36) | 0.49 | 5.57 (2.75) | 5.68 (3.10) | 0.84 |
| **Monocytes** | 10.1 (3.58) | 9.55 (2.58) | 0.20 | 6.92 (3.20) | 7.38 (2.88) | 0.44 |
| **Granulocytes** | 54.7 (11.8) | 55.3 (9.29) | 0.68 | 56.7 (13.0) | 56.5 (12.0) | 0.95 |
| **Nucleated RBC** | 5.04 (6.90) | 5.49 (5.88) | 0.62 | 8.02 (6.81) | 6.87 (4.92) | 0.34 |
| **Mean DNA methylation** |  |  |  |  |  |  |
| **Overall** | 60.9 (0.91) | 60.9 (0.86) | 0.56 | 51.0 (0.63) | 50.9 (0.98) | 0.23 |
| **Open Sea** | 76.7 (1.15) | 76.6 (1.11) | 0.64 | 74.0 (0.80) | 73.7 (1.33) | 0.17 |
| **CpG Shelf** | 78.1 (1.07) | 78.0 (1.01) | 0.69 | 78.4 (0.88) | 78.2 (1.46) | 0.21 |
| **CpG Shore** | 49.6 (0.83) | 49.5 (0.72) | 0.58 | 47.0 (0.65) | 46.8 (0.91) | 0.17 |
| **CpG Island** | 18.0 (0.46) | 17.9 (0.27) | 0.12 | 19.5 (0.53) | 19.4 (0.66) | 0.80 |

Abbreviations: MARBLES, Markers of Autism Risk – Learning Early Signs; EARLI, Early Autism Risk Longitudinal Investigation; RBC, red blood cell

**Table 2.** Distributions of maternal and fetal participant and sample characteristics for those with placenta DNA methylation measures, split by maternal use of prenatal vitamins in month 1 of pregnancy.

|  | MARBLEs |  |  | EARLI |  |  |
| --- | --- | --- | --- | --- | --- | --- |
|  | No use of prenatal vitamins month 1 | Use of prenatal vitamins month 1 | P-value | No use of prenatal vitamins month 1 | Use of prenatal vitamins month 1 | P-value |
|  | N=37 | N=33 |  | N=35 | N=53 |  |
| **Maternal education** |  |  | 0.05 |  |  | 0.33 |
| No Degree | 21 (56.8%) | 10 (30.3%) |  | 17 (48.6%) | 19 (35.8%) |  |
| College Degree | 16 (43.2%) | 23 (69.7%) |  | 18 (51.4%) | 34 (64.2%) |  |
| **Maternal age at delivery (years)** | 35.3 (5.16) | 35.1 (5.86) | 0.89 | 33.8 (4.46) | 33.5 (4.75) | 0.77 |
| **Child race/ethnicity** |  |  | 0.11 |  |  | 0.34 |
| Non-Hispanic White | 16 (43.2%) | 8 (24.2%) |  | 16 (45.7%) | 25 (47.2%) |  |
| Hispanic | 14 (37.8%) | 12 (36.4%) |  | 5 (14.3%) | 11 (20.8%) |  |
| Other/Multiple | 7 (18.9%) | 13 (39.4%) |  | 14 (40.0%) | 17 (32.1%) |  |
| **Child sex** |  |  | 0.05 |  |  | 0.11 |
| Female | 17 (45.9%) | 7 (21.2%) |  | 18 (51.4%) | 17 (32.1%) |  |
| Male | 20 (54.1%) | 26 (78.8%) |  | 17 (48.6%) | 36 (67.9%) |  |
| **Gestational age at delivery (weeks)** | 39.4 (1.14) | 39.2 (0.83) | 0.49 | 39.1 (1.59) | 39.4 (1.22) | 0.39 |
| **Estimated cell proportion** |  |  |  |  |  |  |
| Trophoblasts | 14.0 (7.55) | 15.2 (6.14) | 0.47 | 13.0 (6.04) | 14.4 (5.13) | 0.27 |
| Stromal | 9.06 (6.41) | 10.0 (6.26) | 0.52 | 10.3 (3.75) | 9.47 (3.51) | 0.31 |
| Hofbauer | 4.71 (2.22) | 4.24 (2.22) | 0.37 | 2.73 (1.45) | 2.77 (1.54) | 0.89 |
| Endothelial | 5.43 (2.22) | 7.25 (3.68) | 0.02 | 5.57 (2.45) | 6.05 (2.63) | 0.38 |
| nRBC | 0.80 (1.05) | 0.55 (1.02) | 0.32 | 1.24 (1.00) | 1.46 (2.11) | 0.51 |
| Syncytiotrophoblast | 66.0 (12.6) | 62.8 (9.74) | 0.23 | 67.1 (9.18) | 65.9 (8.63) | 0.51 |
| **Mean DNA methylation** |  |  |  |  |  |  |
| Overall | 53.3 (1.09) | 52.8 (1.12) | 0.05 | 45.0 (1.04) | 44.5 (1.15) | 0.03 |
| Open Sea | 65.3 (1.41) | 64.7 (1.49) | 0.06 | 62.1 (1.17) | 61.6 (1.48) | 0.09 |
| CpG Shelf | 69.2 (1.26) | 68.5 (1.50) | 0.06 | 68.4 (1.29) | 67.8 (1.61) | 0.11 |
| CpG Shore | 45.5 (1.12) | 45.0 (1.09) | 0.06 | 42.7 (1.17) | 42.1 (1.14) | 0.02 |
| CpG Island | 18.7 (0.69) | 18.6 (0.70) | 0.64 | 20.1 (1.00) | 19.6 (0.92) | 0.02 |

### Mean DNA Methylation Differences with Prenatal Vitamin Use

Mean array-wide DNA methylation differed by prenatal vitamin use in the first month of pregnancy in placenta (**Supplemental Figure 3**). In EARLI, prenatal vitamin use was marginally associated with -0.52% (95% confidence interval: -1.04, 0.01) lower mean array-wide placenta DNA methylation. In MARBLES, prenatal vitamin use in the first month was associated with - 0.60% (95% confidence interval: -1.08, -0.13) lower mean array-wide placenta DNA methylation. Differences by prenatal vitamin use were smaller in magnitude for island regions. In cord blood there were no differences in mean array-wide DNA methylation by prenatal vitamin use in either cohort.

### Prenatal Vitamin Epigenome-Wide Association Results

Across all analyses, no single CpG site met the more stringent significance threshold (p-value < 10^−7^). In EARLI placenta tissue 3,442 sites had a nominal association (p-value <0.01). Of these sites, 94.4% had lower DNA methylation with prenatal vitamin use in the first pregnancy month and these sites had an average of -3.9 percent lower DNA methylation (**Figure 1**). The top site associated with prenatal vitamin use in placenta in EARLI was cg24700222 associated with *COL24A1* gene (effect estimate = -7.3, p-value= 2.0×10^−6^) (**Supplemental Table 1**). In MARBLES placental tissue, 9,216 sites had p-value<0.01, and 96.4% had lower DNA methylation with prenatal vitamin use and these sites had an average of - 4.2 percent lower DNA methylation. The top site associated with prenatal vitamin use in placenta in MARBLES was cg00711959 (effect estimate = -11.0, p-value= 3.3×10^−6^), which is located on chromosome 2 and is not annotated to a gene (**Supplemental Table 2**).

**Figure 1.**
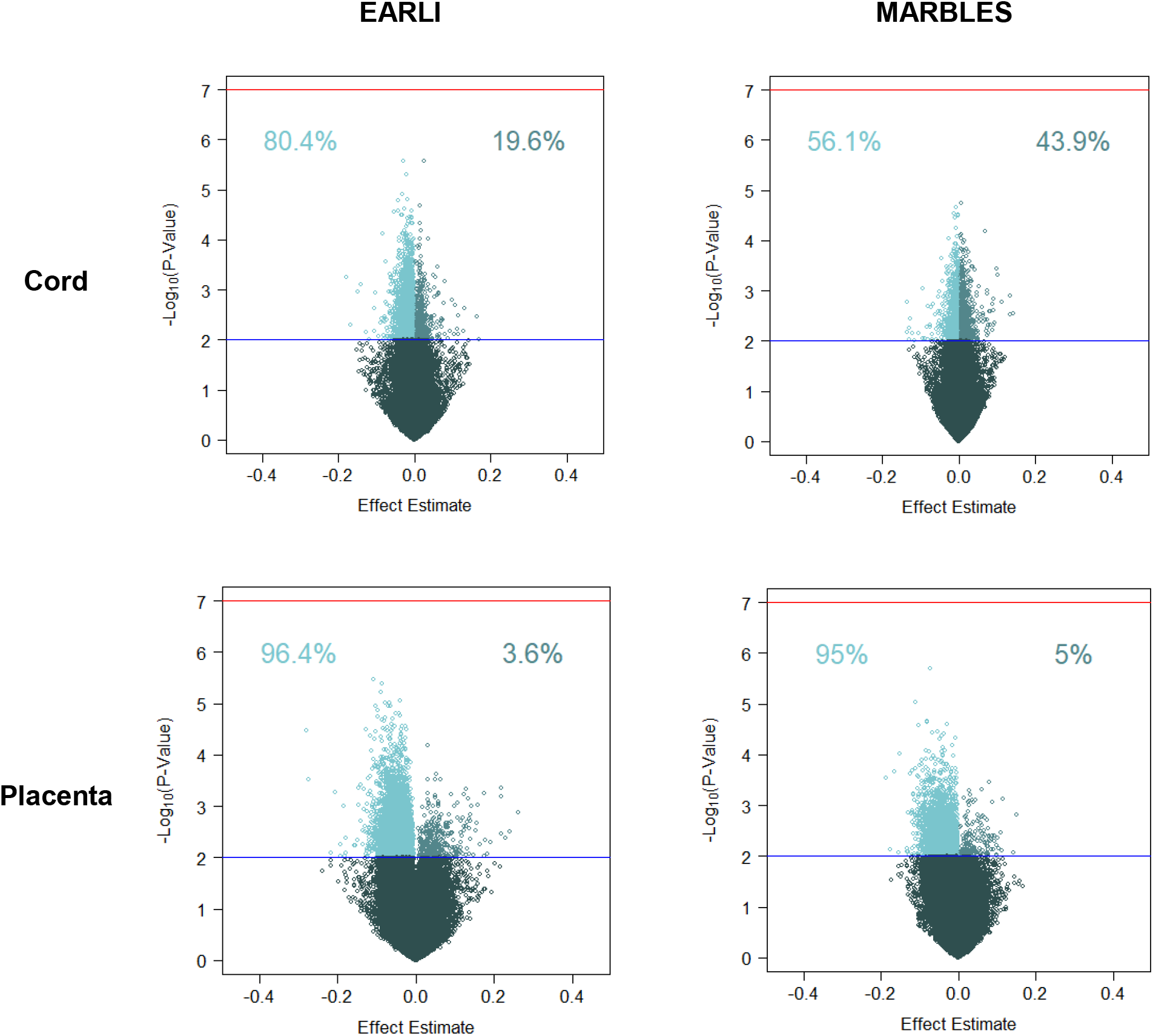
Volcano plots of single site CpG effect estimates for prenatal vitamin use in 1^st^ month of pregnancy and -log_10_(p-values). Percentages indicate proportion of CpG sites with p-value<0.01 that have positive or negative effect estimate. Regression models were adjusted for sex, maternal age, gestational age, maternal education, ancestry PCs, and estimated cell proportions.

In cord blood, among nominally 3,273 associated sites in EARLI, 80.5% had lower DNA methylation with prenatal vitamin use and these sites had an average of 1.8 percent lower DNA methylation. The top site associated with prenatal vitamin use in cord blood in EARLI was cg18452703 (effect estimate = -3.0, p-value= 2.7×10^−6^), located on chromosome 10 and not annotated to a gene. (**Supplemental Table 3**). In MARBLES cord blood, 58.3% of the 2,348 nominally associated sites had lower DNA methylation for prenatal vitamin use and these sites had an average 1.0 percent lower DNA methylation. The top site associated with prenatal vitamin use in cord blood in MARBLES was cg04551619 (effect estimate = 0.56, p-value= 1.8×10^−5^), which is located on chromosome 1 and is not annotated to a gene (**Supplemental Table 4**).

For the DNA methylation sites commonly measured across tissues and cohorts (n=422,383CpGs), overall correlations among effect estimates for the associations of prenatal vitamin use and DNA methylation were weak to modest (**Figure 2**). The strongest correlations in effect estimates were between MARBLES placenta (measured on EPIC) and EARLI placenta (measured on 450k) (r=0.17, p-value<2.2×10^−16^), and between EARLI cord (measured on 450k) and EARLI placenta (r=0.17, p-value<2.2×10^−16^). In contrast, correlation of cord blood effect estimates between the cohorts was non-existent (r=0.02, p-value<2.2×10^−16^), and correlation between cord blood and placenta in MARBLES was similarly weak (r=0.06, p-value<2.2×10^−16^).

**Figure 2.**
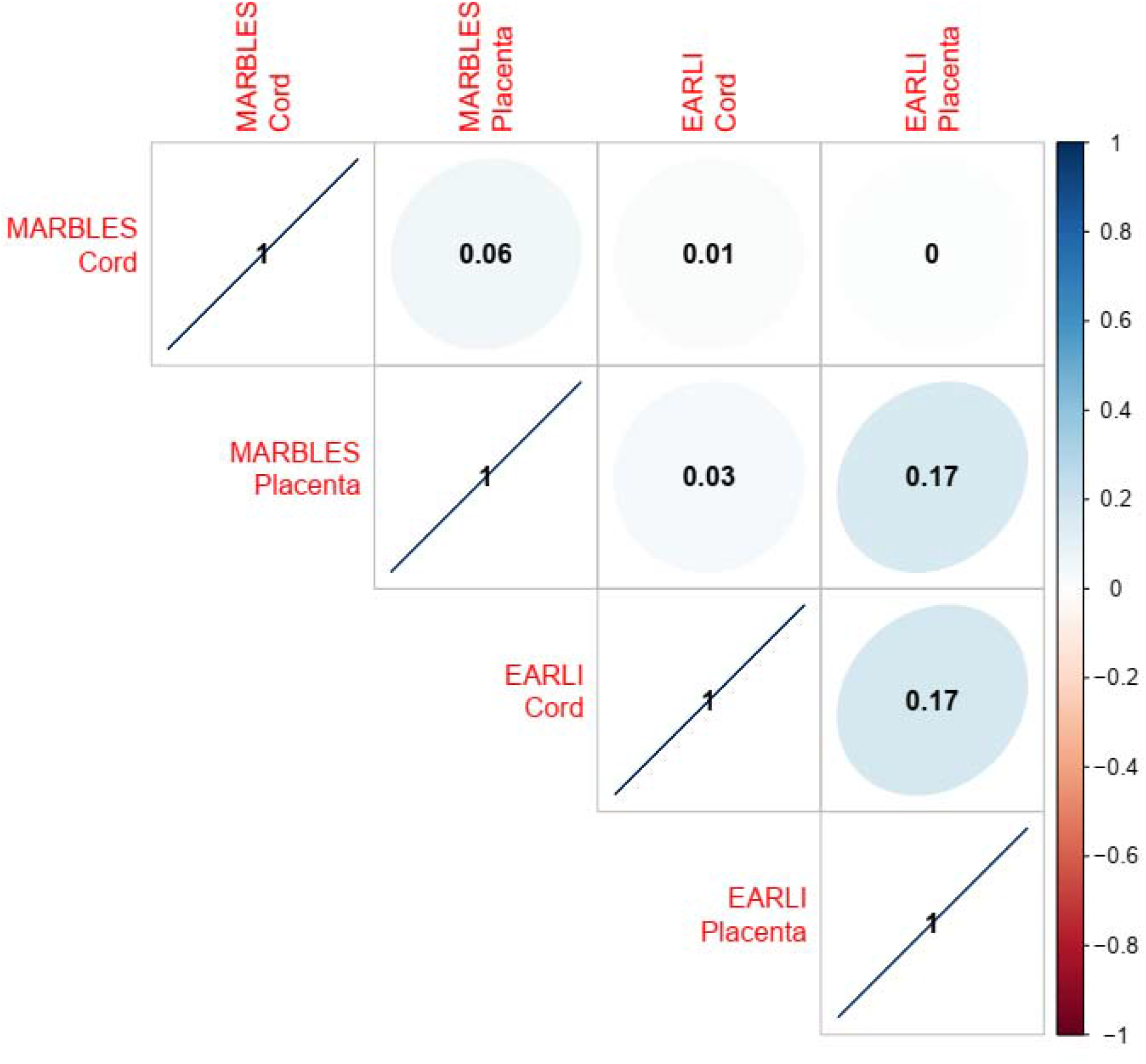
Pearson correlation of regression coefficients across all CpGs in common between EPIC/450k (n=413,011 CpGs).

We restricted to the DNA methylation sites associated (p-value<0.01) with prenatal vitamin use and calculated overlap and correlation across tissues and cohorts. In EARLI cord blood, 4,068 CpGs had p-value<0.01, EARLI placenta had 3,647 CpG sites, MARBLES cord had 4,025 CpGs, and MARBLES placenta had 9,563 CpGs. Sites reaching this threshold were largely tissue and cohort specific (**Figure 3A**). The largest levels of overlap mirrored the correlation patterns. Between EARLI and MARBLES placenta, 101 CpG sites had p-value<0.01, and these 101 sites had correlation r=0.80 (**Figure 3B**). The 101 CpG sites had effect estimates in the same direction in MARBLES and EARLI placenta, except for one site, with 99 of them having negative effect estimates (**Figure 4**). In contrast, comparing cord blood between MARBLES and EARLI there were 20 CpGs with p-value<0.01 in both (**Figure 3B**), with more modest correlation (r=0.50) and mixed directions of effect (**Figure 4**). Between cord blood and placenta in EARLI, 63 sites had p-value<0.01 in both, with those CpG effect estimates having correlation r=0.71. In MARBLES, 31 CpGs had p-value<0.01 in both cord blood and placenta, with effect estimates having correlation r=0.59 (**Figure 3B**). EARLI had more consistency in direction of effect between cord blood and placenta than MARBLES (**Supplemental Figure 4**).

**Figure 3.**
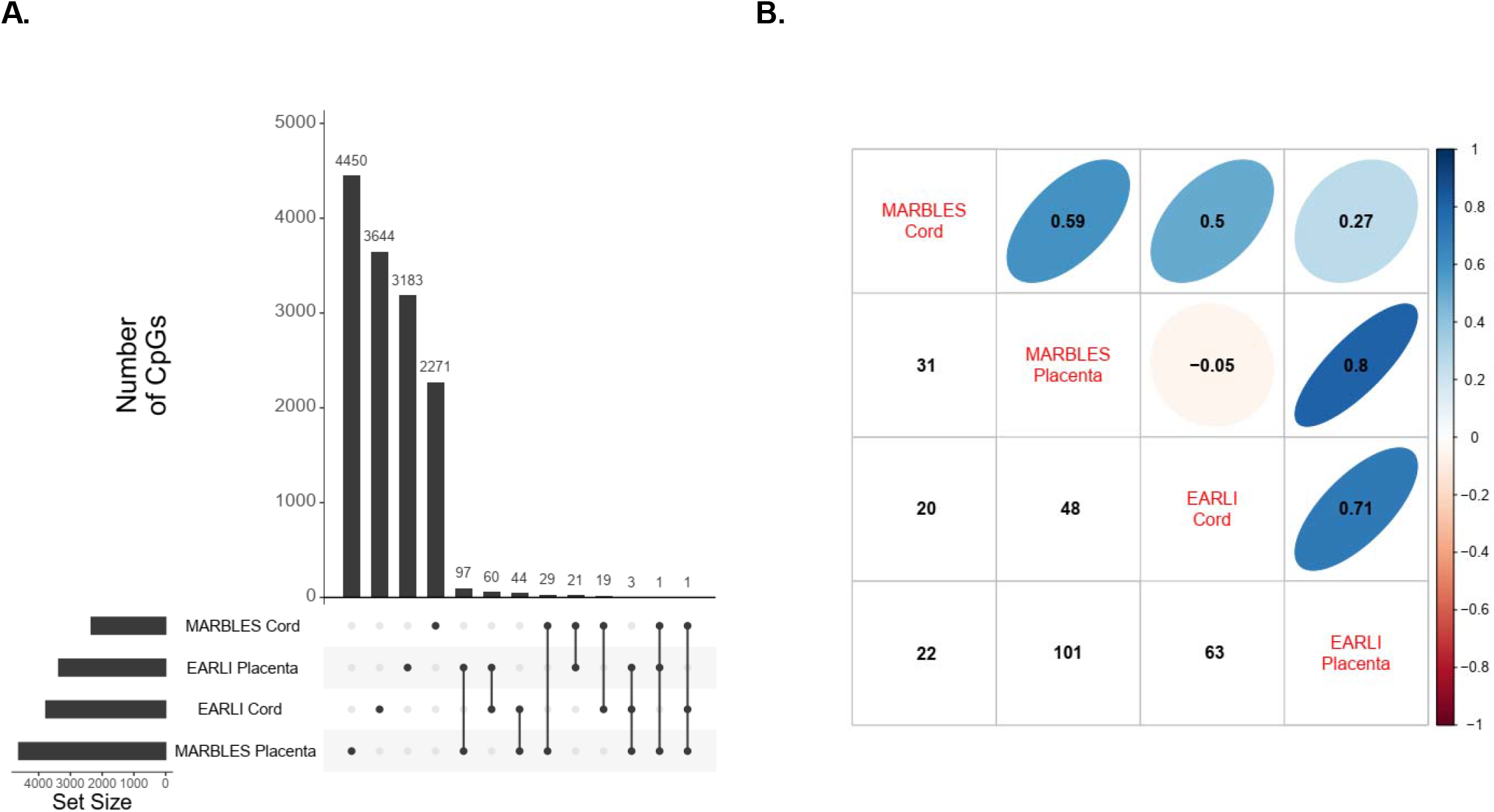
**A)** Number of CpGs with p-value<0.01 unique to and in common with cohorts/tissues. **B)** In upper triangle, correlations between CpG effect estimates with p-value<0.01 in cross comparison, and number of such CpGs shown in lower triangle.

**Figure 4.**
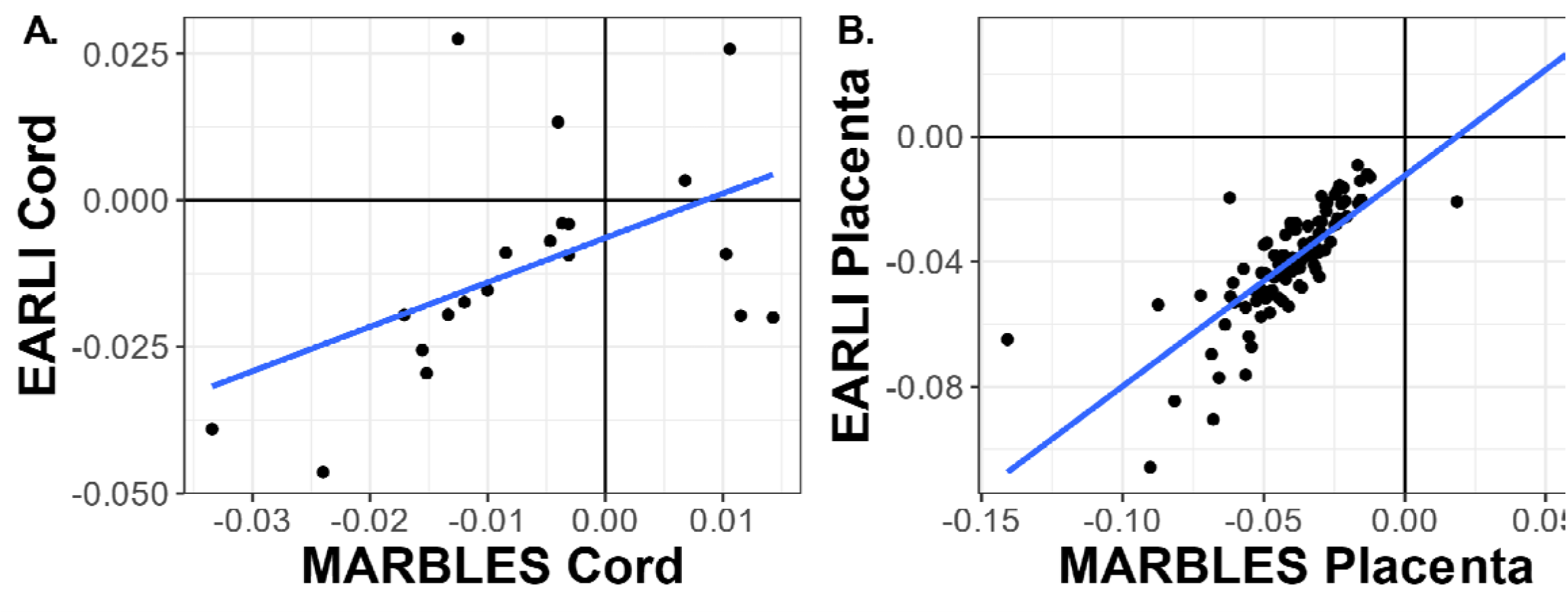
Scatter plots of effect estimates for CpGs with P<0.01 in both cohorts. **A)** Cord blood (n_CpGs_=18) **B)** Placenta (n_CpGs_=101)

We compared single site results with a previous study on maternal plasma folate during pregnancy and cord blood methylation.(16) In that study, 7,219 CpG sites had p-value<0.01. Looking only at sites overlapping between the 450k and EPIC chips, the number of sites with p-value<0.01 was 3,365 in EARLI placenta, 3,771 in EARLI cord, 4,625 in MARBLES placenta, and 2,342 in MARBLES cord. The level of overlap was minimal but more than expected at random between the maternal plasma folate study and our prenatal vitamin usage study, with 72 CpGs overlapping with EARLI placenta results (fisher test p = 0.007), 131 CpGs with EARLI cord (fisher test p < 0.001), 96 CpGs with MARBLES placenta (fisher test p = 0.009), and 45 CpGs with MARBLES cord (fisher test p = 0.10).

### Gene Ontologies Enriched in Prenatal Vitamin Use Related DNA Methylation

CpG sites associated with prenatal vitamin use (p-value<0.01) in the epigenome-wide regression models were used for gene ontology analysis (EARLI cord n_CpGs_ =4,068, EARLI placenta n_CpGs_ =3,647, MARBLES cord n_CpGs_ =4,025, MARBLES placenta n_CpGs_ =9,563). Across all four tissue and cohort combinations, the top pathways by rank sum were neuron and development related pathways, such as nervous system development, neuron differentiation, and cell projection morphogenesis (**Table 3, Supplemental Table 5**). In cord blood, the top pathways by rank sum across the two cohorts included largely development-related pathways, such as sensory organ development, neuron differentiation, and embryo development. In placenta, the top pathways by rank sum across the two cohorts were neuron and synaptic signaling pathways, including neurogenesis, and chemical synaptic transmission.

**Table 3.** Top 20 biologic pathways by rank sum across both cohorts and tissues. Pathway results from missMethyl, using CpG sites associated with prenatal vitamin use in month 1 of pregnancy with P<0.01 as input (MARBLES cord n_CpGs_ =4455, MARBLES placenta n_CpGs_ =9340, EARLI cord n_CpGs_ =4032, EARLI placenta n_CpGs_=3589).

| Biologic pathway | MARBLEs<br>Cord<br>Rank | MARBLEs<br>Placenta<br>Rank | EARLI<br>Cord<br>Rank | EARLI<br>Placenta<br>Rank | Rank<br>Sum |
| --- | --- | --- | --- | --- | --- |
| Behavior | 33 | 29 | 41 | 137 | 240 |
| Nervous system development | 718 | 8 | 1 | 8 | 735 |
| Axon development | 255 | 282 | 368 | 13 | 918 |
| Regulation of synaptic plasticity | 116 | 313 | 122 | 434 | 985 |
| Synapse organization | 760 | 25 | 285 | 42 | 1112 |
| Neurogenesis | 1125 | 5 | 18 | 32 | 1180 |
| Cell morphogenesis involved in differentiation | 424 | 35 | 671 | 121 | 1251 |
| Neuron differentiation | 1218 | 3 | 17 | 39 | 1277 |
| Axonogenesis | 149 | 270 | 884 | 11 | 1314 |
| Neuron development | 1257 | 1 | 178 | 35 | 1471 |
| Central nervous system development | 742 | 67 | 348 | 320 | 1477 |
| Eating behavior | 1069 | 162 | 104 | 260 | 1595 |
| Cognition | 30 | 146 | 197 | 1286 | 1659 |
| Cell morphogenesis involved in neuron differentiation | 663 | 54 | 975 | 46 | 1738 |
| Generation of neurons | 1726 | 10 | 16 | 23 | 1775 |
| Import into cell | 165 | 394 | 513 | 767 | 1839 |
| Neuron recognition | 56 | 716 | 686 | 443 | 1901 |
| Long-term synaptic potentiation | 282 | 363 | 869 | 408 | 1922 |
| Learning or memory | 54 | 334 | 214 | 1376 | 1978 |
| Anatomical structure morphogenesis | 1163 | 471 | 175 | 169 | 1978 |

The top 1000 CpG sites associated with prenatal vitamin use in the first month of pregnancy were examined for enrichment in chromatin state marker signatures. Sites associated with prenatal vitamin use in EARLI cord blood were enriched in the repressed polycomb chromatin state, followed by weak repressed polycomb, bivalent enhancer, and bivalent/poised transcript start sequence markers (**Supplemental Figure 5**).

### Replication Testing with Whole Genome Bisulfite Sequencing (WGBS)

Data for WGBS were available on 63 samples (33 with prenatal vitamin use) in EARLI cord blood, 91 samples (39 with prenatal vitamin use) in MARBLES placenta, 45 samples (17 with prenatal vitamin use) in MARBLES cord on HiSeq 4000, and 42 samples (17 with prenatal vitamin use) in MARBLES cord on HiSeq X Ten. Differentially methylated regions (DMRs) with p-value<0.05 in each are summarized in **Supplemental Table 6**. Overall, a majority of DMRs showed lower DNA methylation with prenatal vitamin use, and agreement in the direction of effect was strongest in placenta, where there were 803 array sites within 5kb of DMRs identified in WGBS, and 66.6% had the same direction of effect across the two measures (**Supplemental Table 7**).

## Discussion

In two prospective pregnancy cohorts we found that prenatal vitamin intake in the first month of pregnancy was related to lower average DNA methylation in placenta and cord blood. The magnitude of this association was strongest in placenta. We observed little consistency in the associations between prenatal vitamin intake and single DNA methylation site effect estimates across cohorts and tissues, with only a few overlapping sites with correlated effect estimates. However, the single DNA methylation sites associated with prenatal vitamin use were consistently enriched in neuron developmental pathways. Together these findings suggest that prenatal vitamin intake may be related to placental global DNA methylation and related to DNA methylation in brain-related pathways in both placenta and cord blood.

Previous research has examined DNA methylation and prenatal vitamin supplementation, or components of prenatal multivitamins, at different supplementation time points. Vitamin B12 is a component of prenatal vitamins, and prenatal deficiency leads to adverse infant hematologic and neurologic outcomes.(28) In terms of DNA methylation, a cohort study of cord blood samples found that higher maternal serum vitamin B12 concentrations (collected at mean 10.6 weeks of gestation) were associated with lower global cord blood DNA methylation, using bisulfite pyrosequencing.(29) Another study of 516 cord blood samples observed periconceptional vitamin B12 intake was not associated with long interspersed nuclear element-1 (LINE-1) DNA methylation, often used as a proxy for global methylation.(30) Placentas from rats who were treated with excess folate and were vitamin B12 deficient had lower global DNA methylation levels compared to controls and rats with normal folate + B12 deficiency, but when they were given omega-3, the DNA methylation returned to the control group level.(31) In humans, mothers with high folate and low vitamin B12 during early pregnancy had significantly lower cord blood DNA methylation as measured by LC-MS/MS.(32) These previous studies were heterogeneous in species, sample type, and DNA methylation measurement methods. The conflicting results highlight the need for more comprehensive analyses of the associations between early pregnancy vitamin supplementation and DNA methylation, considering the intake of multiple vitamins.

Some prenatal vitamins also contain omega-3 fatty acids. The omega-3 fatty acid, docosahexaenoic acid (DHA) and polyunsaturated fatty acids are essential during pregnancy for fetal development.(33) A study of DHA, given at 20+ weeks of gestation, found no difference in global DNA methylation of blood spots and leukocytes between the children from the treatment and control groups.(34) They did identify 21 differentially methylated regions with most regions showing lower methylation levels in the treatment group compared to the control. The direction of this finding matches ours where a majority of our differentially methylated sites (p-value <0.01) across both cohorts and tissue types associated lower methylation with prenatal vitamin use. A randomized control trial of polyunsaturated fatty acid supplementation from weeks 18-22 to birth observed no difference in the cord blood global DNA methylation between the non-smoking treatment and control groups.(35) The review by Andraos *et al* (2018) summarizes the DNA methylation results from randomized control trials of nutritional supplementations during pregnancy, finding overall that micronutrient supplementation does not substantially affect offspring DNA methylation in cord blood, blood spots, placental tissue, and buccal swabs, though lack of standardized methods complicate comparing results of studies.(20)

Folate and folic acid are components of prenatal vitamins. In a randomized control trial of maternal folic acid supplementation in the second and third trimesters, supplementation was associated with lower DNA methylation in cord blood at LINE-1 compared to the control group.(36,37) A meta-analysis of two cohorts of 1,988 infants identified 443 DNA methylation sites in cord blood associated (FDR<0.05) with maternal plasma folate levels measured during pregnancy (median 18 weeks gestation, and median 12.9 weeks gestation in the two cohorts).(16) Of these sites, 416 (94%) had lower DNA methylation with higher plasma folate levels, suggesting a similar direction of association to what we observed. Using gene ontologies, prenatal vitamin-associated DNA methylation sites were enriched for developmental and neuronal pathways in both our cohorts and tissues. The meta-analysis also observed an enrichment of developmental and neurodevelopmental pathways.(16) Although we did not observe correlation of associations at individual DNA methylation sites to this previous study, our findings were consistent with respect to overall trends of lower DNA methylation and pathway enrichment.

Prenatal vitamins overlap with multivitamins in their nutrient components, although prenatal vitamins typically contain more folic acid than multivitamins (about twice as much).(5) Previously in the EARLI cohort, multivitamin usage in the three months before pregnancy in mothers without the *MTHFR* allele variant was associated with higher global cord blood DNA methylation.(19) However, similar to our current findings in cord blood, the previous EARLI study observed no association between prenatal vitamin use in the three months prior to pregnancy and global cord blood DNA methylation (p<0.05). These analyses differed in their exposure window (prior paper: three months prior to pregnancy versus current paper: first month of pregnancy).

Differences in DNA methylation have been associated with other early-life exposures to environmental factors, in addition to prenatal vitamin use.(38–40) In this study, we found larger differences in DNA methylation and more consistency between cohorts in placenta compared to cord blood. This finding is likely due to known DNA methylation characteristics of placenta, including global hypomethylation relative to somatic tissues and the presence of partially methylated domains.(41) In placenta, methylation of gene bodies is predictive of active expression and exposure to home/garden pesticides was previously shown to be associated with higher global DNA methylation in MARBLES placenta samples.(42,43) Combined, these findings suggest that the lower global DNA methylation levels associated with prenatal vitamin use may be reflecting a more quiescent genome with reduced activation of environmentally responsive genes that may be related to the known effects of nutrients in reducing oxidative stress.(44)

While these studies have great heterogeneity with respect to the type, method, and timing of exposure measures, most use the Illumina BeadArrays for DNA methylation measures, though the tissue measured at birth varies. Replication testing or meta-analyses are necessary to determine the reproducibility of these findings and few studies to date have included more than one cohort. These steps will require harmonization of exposure and DNA methylation measures across studies. Once reproducibility and specificity are determined, these DNA methylation patterns may be used as biomarkers of exposure or may predict future health.(22)

There were a number of strengths to this study. We analyzed two prospective pregnancy cohorts and two tissue types with 201 total samples from EARLI and 271 total samples from MARBLES. The prospective design minimized recall bias of prenatal vitamin use. Furthermore, we had consistent exposure measures including using data from questionnaires, consistent cleaning of the vitamin questionnaires, and consistent exposure timing during pregnancy. The results were based on a diverse subset of participants from five study sites in the US. We employed rigorous methods to preprocess and analyze the DNA methylation data. Our analysis across the two cohorts and two tissue types showed similar gene ontology pathways. Finally, we compared the array-based DNA methylation single site results to the WGBS results.

There were several limitations to our study. First, the samples used for DNA methylation were from one time point at birth, so long-term differences in DNA methylation were not assessed. We examined prenatal vitamin use as a yes/no response for any use in the first pregnancy month; effects may differ by frequency of intake, nutrient composition, and nutrient dose. We did not account for differences in underlying nutritional deficiencies which may have affected our results. Future studies could assess nutritional intake in addition to prenatal vitamins. Exposure to other environmental chemicals may lower the DNA methylation levels of adults, children, and infants.(29,45–49) It is possible that differences in chemical exposures may account for or mask some of the differences across DNA methylation related to prenatal vitamin use. While there was little positional overlap in effects between array-based DNA methylation and WGBS, we observed similar directions of effect across measurement methods in placenta. This finding may have been due to a greater overlap in these samples between measured on array compared to WGBS. Data on method of delivery, known to affect DNA methylation,(50) were incomplete in EARLI, so we did not adjust for that information in our regression analyses for both cohorts. We observed a higher correlation between EARLI cord and placenta that may have been a result of the DNA methylation being measured at the same time (**Supplemental Figure 4**). MARBLES cord blood and placenta DNA methylation were measured in two separate batches, so batch differences and tissue differences are correlated. Future studies could examine a larger cohort, compare these results to a general population cohort, or conduct a meta-analysis of multiple prenatal vitamin studies. Future studies may also consider examining the placenta, to explore whether it is more sensitive to effects of supplementation, as our study suggests.

## Conclusions

We found that prenatal vitamin use in the first month of pregnancy was associated with lower DNA methylation, particularly in the placenta. Prenatal vitamin use is recommended before and during pregnancies for normal fetal development. Given its importance, additional research is needed to understand the underlying biological mechanisms of development. By demonstrating an association between prenatal vitamin intake and DNA methylation at birth, we lay the foundation for DNA methylation as a biomarker of prenatal vitamin exposure.

## Methods

### Study samples

The Early Autism Risk Longitudinal Investigation (EARLI) and Markers of Autism Risk Learning Early Signs (MARBLES) studies are enriched risk prospective pregnancy cohorts studying autism etiology.(51,52) The EARLI study was reviewed and approved by Human Subjects Institutional Review Boards (IRBs) from each of the four study sites (Johns Hopkins University, Drexel University, University of California Davis, and Kaiser Permanente Northern California). The MARBLES protocol was reviewed and approved by the Human Subjects IRB from University of California Davis. Secondary data analysis for this manuscript was approved by the Human Subjects IRB for the University of Michigan. Both studies recruited mothers of children with clinically confirmed ASD who were early in a subsequent pregnancy or were trying to become pregnant. In EARLI there were 232 mothers with a subsequent sibling born through this study between November 2009 and March 2012. In MARBLES there were 389 enrolled mothers that gave birth to 425 subsequent siblings between December 1, 2006 and July 1, 2016.

### Covariate and exposure assessment

Demographics, behaviors, and medical history were all collected via maternal self-report questionnaire. In these questionnaires, mothers were asked if they used prenatal vitamins for each month of pregnancy (yes/no). Data for the first month of pregnancy was collected at study enrollment.

### Sample Collection and Processing

In EARLI, biospecimens including cord blood and placenta, were collected and archived for 213 births. Full thickness placental tissue from a central cotyledon was collected. Sterile punch biopsy forceps were used to extract placental samples from the maternal and fetal sides. Whole cord blood was also collected at delivery. Samples were transported to the Johns Hopkins Biological Repository (JHBR) for aliquoting and archiving (−80ºC). Placental DNA was extracted with the DNeasy Tissue Kit (Qiagen), and cord blood DNA was extracted using the DNA Midi kit (Qiagen, Valencia, CA). DNA was quantified using the Nanodrop (ThermoFisher Waltham, MA) and normalized DNA aliquots were sent to the Center for Inherited Disease Research (Johns Hopkins University). DNA samples were bisulfite treated and cleaned using the EZ DNA methylation gold kit (Zymo Research, Irvine, CA) according to manufacturer’s instructions. DNA was plated randomly and was assayed on the Infinium HumanMethylation450 BeadChip (Illumina, San Diego, CA).(53) Methylation control gradients and between-plate repeated tissue controls were used.

In MARBLES, placental tissues and cord blood were collected at delivery and immediately processed and frozen. The MARBLES study used orientation to the umbilical cord to ensure that all placenta samples were isolated from the chorionic villus from the fetal side of the placenta. Placental and cord blood samples were stored at -80ºC in the UC Davis repository. Cord blood and placenta samples were processed for methylation measures. Placenta DNA was extracted with Gentra Puregene kit (Qiagen) and cord blood DNA was extracted using the DNA Midi kit (Qiagen, Valencia, CA). Samples were bisulfite treated and cleaned using the EZ DNA methylation gold kit (Zymo Research, Irvine, CA). DNA was plated randomly and assayed on the Infinium HumanMethylationEPIC BeadChip (Illumina, San Diego, CA) at the Johns Hopkins SNP Center, a shared lab and informatics operation with the Center for Inherited Disease Research (Johns Hopkins University). DNA methylation control gradients and between-plate repeated tissue controls were used.

### DNA Methylation Processing

For all methylation samples, we used the minfi library (version 1.30.0) in R (version 3.6) to process raw Illumina image files into noob background corrected methylation values.(54,55) In EARLI, cord blood and placenta samples were run on the 450k array together, and thus preprocessed together. Samples from multiple births (cord blood n=2 samples, placenta n=6 samples), as well as samples with discordant DNA methylation predicted sex and observed infant sex were removed (cord blood n=3, placenta n=1). Probes with failed detection P-value (>0.01) in >5% of samples were removed (n=661), as were probes documented as cross reactive (n=29,153).(56) Y-chromosome probes (n=48) were dropped from analysis. There were 170 EARLI cord blood samples, and 127 EARLI placenta samples with 455,650 probes that passed DNA methylation quality control.

In MARBLES, placenta and cord blood samples were run on the EPIC array at different times and preprocessed separately. First, we dropped cord blood samples from multiple births (cord blood n=8 samples). Samples that had mismatched predicted sex were dropped (cord n=3). For siblings not from multiple births, all but one sibling was dropped (cord n=13). Probes were dropped if they had detection-p (p>0.01) failure in greater than 5% of samples (n=4,630). Cross reactive probes (n=42,967) and Y chromosome probes were dropped from analysis (n=379).(57) There were 243 MARBLES cord blood samples with 817,883 probes that passed DNA methylation quality control. Second, no placenta samples had mismatched predicted sex. There were no samples from multiple births, and all but one sample from siblings were dropped (placenta n=2). Probes that failed detection-p in >5% of samples (n=1,699), cross reactive probes (n=43,068), and remaining Y-chromosome probes were dropped from analysis (n=84). There were 90 MARBLES placenta samples with 821,008 probes that passed quality control. Sample exclusion is summarized in **Supplemental Figure 1** and CpG probe exclusion is summarized in **Supplemental Figure 2**.

In cord blood samples, cell type (CD8^+^ T-cell, CD4^+^ T-cell, natural killer cell, B-cell, monocyte, granulocyte, and nucleated red blood cell) proportions were estimated using a combined reference panel with the IDOL method.(58) In placenta samples, EpiDISH(59) was used to predict proportions of placenta cell types using a reference panel from the planet package: trophoblasts, stromal cells, Hofbauer cells, endothelial cells, nucleated red blood cells, and syncytiotrophoblasts.(60) Mean DNA methylation per person was calculated as the mean across all probes.(19) Mean DNA methylation restricted to probes in genomic regions (CpG island, shore, shelf, or open sea) were also computed. We used Illumina’s annotation of CpG sites to assign genomic regions (CpG island, CpG shore, CpG shelf, open sea).(61,62)

### Genetics Data Processing

In EARLI, genetic data were measured using the Omni5+exome array (Illumina) at the John Hopkins University Center of Inherited Disease Research (CIDR). Data on 4.6 million single nucleotide polymorphisms (SNPs) were generated for 841 EARLI family biosamples (including maternal, paternal, proband, and infant samples) from 254 families and 18 HapMap control samples. Samples were processed together, but only data from infants with cord blood or placenta methylation were used. No samples had missing genotypes at >3% of probes, or excess heterozygosity or homozygosity (4 standard deviations). Probes were removed if they had technical problems flagged by CIDR or missing genomic location information. Single nucleotide polymorphisms (SNPs) with minor allele frequencies >5% were removed if they had a missingness rate >5%, and SNPs with minor allele frequency <5% were removed if they had a missingness rate >1%. There were 2.5 million clean SNPs for 827 samples, which were merged with the 1000 genomes project (1000GP, version 5) data(63) and principal components for genetic ancestry were computed.

In MARBLES, SNPs on 643 infant and mother samples from 234 families were genotyped using the Illumina Mega array at the John Hopkins University Center of Inherited Disease Research (CIDR). Maternal and infant samples were processed together, but only data from infants with cord blood or placenta methylation measures were used. We again applied stringent quality control criteria(64) to the raw 1.75 million genotypes to remove low quality SNPs and samples. Our criteria include removal of samples with call rate <98%, sex discrepancy, and relatedness (pi-hat < 0.18) to non-familial samples. We also filtered SNPs with call rates < 95%, excess hetero-or homozygosity, and minor allele frequency (MAF) < 5%. After quality control, 620 samples and 758 thousand SNPs remained. Principal components were calculated on genotype data, and these principal components were used to adjust for genetic ancestry in models.

### Statistical Analyses

Study sample descriptive statistics were calculated for each of the four cohort/tissue groups. For continuous covariates (maternal age at delivery, gestational age, estimated cell proportions), we calculated mean and standard deviation. For categorical covariates (maternal education, infant sex, infant race/ethnicity), we provided number and frequency. We calculated the bivariate relationships between covariates and prenatal vitamin use. For continuous covariates, we used t-tests and for categorical covariates, we used chi-square tests.

In multivariable linear regression analyses, first, we examined array wide mean DNA methylation differences by prenatal vitamin intake in the first month of pregnancy. Regression models were adjusted for infant sex, maternal age, gestational age, maternal education, genetic ancestry principal components, and estimated cell proportions. Since cell composition estimates sum up to 100%, to avoid collinearity issues in models, we did not use all predicted cell types in models. For placenta, syncytiotrophoblast and Hofbauer proportions were used, while in cord blood granulocyte and nucleated red blood cell proportions were used. We visualized regression coefficients and 95% confidence intervals using forest plots.

Next, we performed epigenome-wide association analyses by examining single CpG site differential DNA methylation. We fit parallel linear models for each probe. Models were again adjusted for infant sex, maternal age, gestational age, maternal education, genetic ancestry PCs, and estimated cell proportions. Regression and empirical Bayes standard error moderation were performed using the limma package.(65) We visualized findings using volcano plots of effect estimates and -log10(p-values). For sites reaching a nominal p-value threshold (p<0.05), we calculated the proportion of sites that had higher DNA methylation with prenatal vitamin intake and the proportion of sites with lower DNA methylation. To compare pairwise results across cohort and tissues, we examined Pearson correlations of effect estimates from these regression models, across all sites in common between the 450k and EPIC methylation arrays. We also focused on CpG sites that had p-value<0.01 in multiple cohort/tissues, examining the overlap of such sites with an upset plot and the Pearson correlation of overlapping sites. For sites prioritized in multiple cohorts/tissues, we also used scatter plots to visualize the effect estimates.

We tested enrichment for gene ontology biological processes using the missMethyl package.(66) As input to missMethyl, CpG sites with p-value<0.01 in the epigenome-wide regression models were used. We ranked the gene ontologies by significance, then computed a rank sum by adding the ranks across the four cohort/tissue groups. In addition, we tested for enrichment of chromatin state types using eFORGE 2.0.(67) The top 1000 CpG sites for each cohort/tissue analysis was input into the eFORGE site, with appropriate array platform chosen (450k for EARLI, EPIC for MARBLES), and Consolidated Roadmap Epigenomics – All 15-state marks and 1 kb window proximity options, with other options set at defaults.

### Replication Testing

In EARLI, whole genome bisulfite sequencing (WGBS) data were available on 63 cord blood samples sequenced on the HiSeq X (51 overlapping with array methylation data). Sample processing and WGBS quality control and alignment for cord blood samples(68) and placenta samples(69,70) have been previously discussed. In MARBLES, WGBS data were available for 91 placenta samples sequenced on HiSeq X Ten (89 overlapping with the array methylation data), 45 cord blood samples sequenced on HiSeq 4000 (30 overlapping with array methylation data), and 42 cord blood samples sequenced on the HiSeq X (35 overlapping with array methylation data).

Raw sequencing reads were preprocessed, mapped to human genome, and converted to CpG methylation count matrices with CpG_Me using default parameters(71–73). Reads were trimmed for adapters and methylation bias, aligned to the reference genome, and filtered for PCR duplicates. Methylation counts at all sites were extracted to Bismark cytosine methylation reports. The CpG_Me workflow incorporates Trim Galore, Bismark, Bowtie2, SAMtools, and MultiQC.(72,74–77)

Differentially methylated regions (DMRs) were identified between prenatal vitamin intake during the first month of pregnancy with adjustment for sex and 10 permutation tests using DMRichR.(71) The DMR analysis utilizes a smoothing and weighting algorithm to weight regions with high coverage and low variation. Permutation testing was performed on pooled null distribution to generate empirical p-values as significant DMRs. The DMRichR pipeline utilized dmrseq and bsseq algorithms.(78,79) To evaluate consistency with array results, we identified array probes within 5kb of the DMR and examined concordance in estimated directions of effect of those CpG probes and the DMR.

## Data Availability

Data are available through the National Database for Autism Research (NDAR) accession numbers for EARLI (1600) and MARBLES (2462).

## Abbreviations

EARLI: Early Autism Risk Longitudinal Investigation
DHA: Docosahexaenoic acid
DMRs: Differentially Methylated Regions
LINE-1: Long Interspersed Nuclear Element-1
MARBLES: Markers of Autism Risk Learning Early Signs

## Declarations

### Ethics approval and consent to participate

The EARLI study was reviewed and approved by Human Subjects Institutional Review Boards (IRBs) from each of the four study sites (Johns Hopkins University, Drexel University, University of California Davis, and Kaiser Permanente Northern California). The MARBLES protocol was reviewed and approved by the Human Subjects IRB from University of California Davis. The University of Michigan Institutional Review Board (HUM00116291) approved these secondary data analyses.

### Consent for publication

All co-authors read and approved the final manuscript.

### Availability of data and materials

Data are available through the National Database for Autism Research (NDAR) accession numbers for EARLI (1600) and MARBLES (2462). Code files are available through GitHub (https://github.com/bakulskilab).

### Competing interests

Not applicable

### Funding

Nutrient and DNA methylation measures and analyses were supported by the National Institutes of Health (R01 ES025574, PI: Schmidt). Cord blood DNA methylation was also funded by R01 ES025531, PI: Fallin. Funding for the EARLI study was provided by the National Institutes of Health (R01ES016443, PI: Newschaffer) and Autism Speaks (003953 PI: Newschaffer). The MARBLES study was funded by National Institutes of Health grants (R24ES028533, R01ES020392, R01ES028089, R01ES020392, P01ES011269) and an EPA STAR grant (#RD-83329201). Support for this research was also provided by the National Institutes of Health grants (P30 ES017885 and R24 ES030893, PI: Fallin). Mr. Dou and Dr. Bakulski were also supported by grants (R01 ES025531, PI: Fallin; R01 AG067592, MPI: Bakulski; R01 MD013299). Dr. Zhu and Dr. LaSalle were supported by R01 ES029213. The content is solely the responsibility of the authors and does not necessarily represent the official views of the National Institutes of Health.

### Authors’ contributions

JD compiled and analyzed the data from the EARLI and MARBLES cohorts and was a major contributor in writing the manuscript.

LM was a major contributor in writing the manuscript.

YZ generated and analyzed the placenta whole genome bisulfite sequencing data

KB analyzed the genetics data

JF generated and curated data

LC developed the cohort and provided manuscript review and editing.

IH-P developed the cohort and provided manuscript review and editing.

CN developed the cohort and provided manuscript review and editing.

JL provided supervision, and provided manuscript review and editing.

DF developed the cohort and provided manuscript review and editing.

RS provided funding and project conceptualization and project management, manuscript review and editing.

KB conceptualized the manuscript and contributed to the original draft and revisions.

All co-authors read and approved the final manuscript.

## Acknowledgements

(Not applicable)

## Notes

### Competing Interest Statement

The authors have declared no competing interest.

### Author Declarations

Secondary data analysis for this manuscript was approved by the Human Subjects IRB for the University of Michigan.

## References Cited

1. World Health Organization (Organization). Essential Nutrition Actions: improving maternal, newborn, infant and young child health and nutrition. Geneva; 2013.

2. The American College of Obstetricians and Gynecologists. Nutrition During Pregnancy [Internet]. 2021 [cited 2021 Mar 9]. Available from: https://www.acog.org/womens-health/faqs/nutrition-during-pregnancy?utm_source=redirect&utm_medium=web&utm_campaign=int#:~:text=When you are pregnant you, first 12 weeks of pregnancy

3. Ottney A, Lebeau L. A “secret shopper” survey of community pharmacist prenatal vitamin recommendations. Journal of the American Pharmacists Association. 2021;

4. Duerbeck NB, Dowling DD, Duerbeck JM. Prenatal Vitamins: What Is in the Bottle? Obstetrical & Gynecological Survey. 2014;69(12).

5. Schmidt RJ, Hansen RL, Hartiala J, Allayee H, Schmidt LC, Tancredi DJ, et al. Prenatal vitamins, one-carbon metabolism gene variants, and risk for autism. Epidemiology (Cambridge, Mass). 2011 Jul;22(4):476–85.

6. Brieger KK, Bakulski KM, Pearce CL, Baylin A, Dou JF, Feinberg JI, et al. The Association of Prenatal Vitamins and Folic Acid Supplement Intake with Odds of Autism Spectrum Disorder in a High-Risk Sibling Cohort, the Early Autism Risk Longitudinal Investigation (EARLI). Journal of Autism and Developmental Disorders. 2021;

7. Branum AM, Bailey R, Singer BJ. Dietary Supplement Use and Folate Status during Pregnancy in the United States. The Journal of Nutrition. 2013 Apr 1;143(4):486–92.

8. Aronsson CA, Vehik K, Yang J, Uusitalo U, Hay K, Joslowski G, et al. Use of dietary supplements in pregnant women in relation to sociodemographic factors – a report from The Environmental Determinants of Diabetes in the Young (TEDDY) study. Public Health Nutrition. 2013/03/04 ed. 2013;16(8):1390–402.

9. Sullivan KM, Ford ES, Azrak MF, Mokdad AH. Multivitamin Use in Pregnant and Nonpregnant Women: Results from the Behavioral Risk Factor Surveillance System. Public Health Reports. 2009 May 1;124(3):384–90.

10. Saldanha LG, Dwyer JT, Andrews KW, Brown LL, Costello RB, Ershow AG, et al. Is Nutrient Content and Other Label Information for Prescription Prenatal Supplements Different from Nonprescription Products? Journal of the Academy of Nutrition and Dietetics. 2017;117(9):1429–36.

11. Bailey RL, Pac SG, Fulgoni III VL, Reidy KC, Catalano PM. Estimation of Total Usual Dietary Intakes of Pregnant Women in the United States. JAMA Network Open. 2019 Jun 21;2(6):e195967–e195967.

12. Sfakianaki MD, Mph AK. Prenatal vitamins: A review of the literature on benefits and risks of various nutrient supplements. Formulary. 2013 Feb;48(2):77–82.

13. Patel A, Lee SY, Stagnaro-Green A, MacKay D, Wong AW, Pearce EN. Iodine Content of the Best-Selling United States Adult and Prenatal Multivitamin Preparations. Thyroid. 2018 Sep 28;29(1):124–7.

14. Vahdaninia M, Mackenzie H, Helps S, Dean T. Prenatal Intake of Vitamins and Allergic Outcomes in the Offspring: A Systematic Review and Meta-Analysis. The journal of allergy and clinical immunology In practice. 2017;5(3):771-778.e5.

15. Hoyo C, Murtha AP, Schildkraut JM, Jirtle RL, Demark-Wahnefried W, Forman MR, et al. Methylation variation at IGF2 differentially methylated regions and maternal folic acid use before and during pregnancy. Epigenetics. 2011/07/01 ed. 2011 Jul;6(7):928–36.

16. Joubert BR, den Dekker HT, Felix JF, Bohlin J, Ligthart S, Beckett E, et al. Maternal plasma folate impacts differential DNA methylation in an epigenome-wide meta-analysis of newborns. Nature Communications. 2016;7(1):10577.

17. Suderman M, Stene LC, Bohlin J, Page CM, Holvik K, Parr CL, et al. 25-Hydroxyvitamin D in pregnancy and genome wide cord blood DNA methylation in two pregnancy cohorts (MoBa and ALSPAC). The Journal of steroid biochemistry and molecular biology. 2016/03/04 ed. 2016 May;159:102–9.

18. Lecorguillé M, Charles M-A, Lepeule J, Lioret S, de Lauzon-Guillain B, Forhan A, et al. Association between dietary patterns reflecting one-carbon metabolism nutrients intake before pregnancy and placental DNA methylation. Epigenetics. 2021 Aug 31;1–16.

19. Bakulski KM, Dou JF, Feinberg JI, Brieger KK, Croen LA, Hertz-Picciotto I, et al. Prenatal Multivitamin Use and MTHFR Genotype Are Associated with Newborn Cord Blood DNA Methylation. Vol. 17, International Journal of Environmental Research and Public Health. 2020.

20. Andraos S, de Seymour JV, O’Sullivan JM, Kussmann M. The Impact of Nutritional Interventions in Pregnant Women on DNA Methylation Patterns of the Offspring: A Systematic Review. Molecular Nutrition & Food Research. 2018 Dec 1;62(24):1800034.

21. Robertson KD. DNA methylation and human disease. Vol. 6, Nature Reviews Genetics. 2005. p. 597–610.

22. James P, Sajjadi S, Tomar AS, Saffari A, Fall CHD, Prentice AM, et al. Candidate genes linking maternal nutrient exposure to offspring health via DNA methylation: a review of existing evidence in humans with specific focus on one-carbon metabolism. International Journal of Epidemiology. 2018 Dec 1;47(6):1910–37.

23. Spurway J, Logan P, Pak S. The development, structure and blood flow within the umbilical cord with particular reference to the venous system. Australasian Journal of Ultrasound in Medicine. 2012 Aug 1;15(3):97–102.

24. Broxmeyer HE, Douglas GW, Hangoc G, Cooper S, Bard J, English D, et al. Human umbilical cord blood as a potential source of transplantable hematopoietic stem/progenitor cells. Proceedings of the National Academy of Sciences. 1989 May 1;86(10):3828 LP – 3832.

25. Bakulski KM, Feinberg JI, Andrews S V., Yang J, Brown S, L. McKenney S, et al. DNA methylation of cord blood cell types: Applications for mixed cell birth studies. Epigenetics. 2016 May 3;11(5):354–62.

26. Gude NM, Roberts CT, Kalionis B, King RG. Growth and function of the normal human placenta. Thrombosis Research. 2004;114(5):397–407.

27. Kaufmann P, Frank H-G. Chapter 10 - Placental Development. In: Polin RA, Fox WW, Abman Shbt-f and NP (Third E, editors. W.B. Saunders; 2004. p. 85–97.

28. Finkelstein JL, Layden AJ, Stover PJ. Vitamin B-12 and Perinatal Health. Advances in Nutrition. 2015 Sep 1;6(5):552–63.

29. McKay JA, Groom A, Potter C, Coneyworth LJ, Ford D, Mathers JC, et al. Genetic and non-genetic influences during pregnancy on infant global and site specific DNA methylation: role for folate gene variants and vitamin B12. PloS one. 2012/03/30 ed. 2012;7(3):e33290– e33290.

30. Boeke CE, Baccarelli A, Kleinman KP, Burris HH, Litonjua AA, Rifas-Shiman SL, et al. Gestational intake of methyl donors and global LINE-1 DNA methylation in maternal and cord blood: Prospective results from a folate-replete population. Epigenetics. 2012 Mar 1;7(3):253–60.

31. Kulkarni A, Dangat K, Kale A, Sable P, Chavan-Gautam P, Joshi S. Effects of Altered Maternal Folic Acid, Vitamin B12 and Docosahexaenoic Acid on Placental Global DNA Methylation Patterns in Wistar Rats. PLOS ONE. 2011 Mar 10;6(3):e17706.

32. Plumptre L, Tammen SA, Sohn K-J, Masih SP, Visentin CE, Aufreiter S, et al. Maternal and Cord Blood Folate Concentrations Are Inversely Associated with Fetal DNA Hydroxymethylation, but Not DNA Methylation, in a Cohort of Pregnant Canadian Women. The Journal of Nutrition. 2020 Feb 1;150(2):202–11.

33. Coletta JM, Bell SJ, Roman AS. Omega-3 Fatty acids and pregnancy. Reviews in obstetrics & gynecology. 2010;3(4):163–71.

34. van Dijk SJ, Zhou J, Peters TJ, Buckley M, Sutcliffe B, Oytam Y, et al. Effect of prenatal DHA supplementation on the infant epigenome: results from a randomized controlled trial. Clinical Epigenetics. 2016;8(1):114.

35. Lee H-S, Barraza-Villarreal A, Hernandez-Vargas H, Sly PD, Biessy C, Ramakrishnan U, et al. Modulation of DNA methylation states and infant immune system by dietary supplementation with ω-3 PUFA during pregnancy in an intervention study. The American Journal of Clinical Nutrition. 2013 Aug 1;98(2):480–7.

36. Caffrey A, Irwin RE, McNulty H, Strain JJ, Lees-Murdock DJ, McNulty BA, et al. Gene-specific DNA methylation in newborns in response to folic acid supplementation during the second and third trimesters of pregnancy: epigenetic analysis from a randomized controlled trial. The American journal of clinical nutrition. 2018 Apr;107(4):566–75.

37. Yang AS, Estécio MRH, Doshi K, Kondo Y, Tajara EH, Issa JJ. A simple method for estimating global DNA methylation using bisulfite PCR of repetitive DNA elements. Nucleic Acids Research. 2004 Feb 1;32(3):e38–e38.

38. Sen A, Heredia N, Senut M-C, Hess M, Land S, Qu W, et al. Early life lead exposure causes gender-specific changes in the DNA methylation profile of DNA extracted from dried blood spots. Epigenomics. 2015 Jun 1;7(3):379–93.

39. Demetriou CA, van Veldhoven K, Relton C, Stringhini S, Kyriacou K, Vineis P. Biological embedding of early-life exposures and disease risk in humans: a role for DNA methylation. European Journal of Clinical Investigation. 2015 Mar 1;45(3):303–32.

40. Mitchell C, Schneper LM, Notterman DA. DNA methylation, early life environment, and health outcomes. Pediatric Research. 2016;79(1):212–9.

41. Schroeder DI, Blair JD, Lott P, Yu HOK, Hong D, Crary F, et al. The human placenta methylome. Proceedings of the National Academy of Sciences of the United States of America. 2013 Apr;110(15):6037–42.

42. Schroeder DI, Jayashankar K, Douglas KC, Thirkill TL, York D, Dickinson PJ, et al. Early Developmental and Evolutionary Origins of Gene Body DNA Methylation Patterns in Mammalian Placentas. PLoS genetics. 2015 Aug 4;11(8):e1005442–e1005442.

43. Schmidt RJ, Schroeder DI, Crary-Dooley FK, Barkoski JM, Tancredi DJ, Walker CK, et al. Self-reported pregnancy exposures and placental DNA methylation in the MARBLES prospective autism sibling study. Environmental Epigenetics. 2016 Dec 1;2(4).

44. Rodríguez-Cano AM, Calzada-Mendoza CC, Estrada-Gutierrez G, Mendoza-Ortega JA, Perichart-Perera O. Nutrients, Mitochondrial Function, and Perinatal Health. Nutrients. 2020 Jul 21;12(7):2166.

45. Rusiecki JA, Andrea B, Valentina B, Letizia T, Moore LE, Bonefeld-Jorgensen EC. Global DNA Hypomethylation Is Associated with High Serum-Persistent Organic Pollutants in Greenlandic Inuit. Environmental Health Perspectives. 2008 Nov 1;116(11):1547–52.

46. Keon-Yeop K, Dong-Sun K, Sung-Kook L, In-Kyu L, Jung-Ho K, Yoon-Seok C, et al. Association of Low-Dose Exposure to Persistent Organic Pollutants with Global DNA Hypomethylation in Healthy Koreans. Environmental Health Perspectives. 2010 Mar 1;118(3):370–4.

47. Bollati V, Baccarelli A, Hou L, Bonzini M, Fustinoni S, Cavallo D, et al. Changes in DNA Methylation Patterns in Subjects Exposed to Low-Dose Benzene. Cancer Research. 2007 Feb 1;67(3):876 LP – 880.

48. Baccarelli A, Wright RO, Bollati V, Tarantini L, Litonjua AA, Suh HH, et al. Rapid DNA methylation changes after exposure to traffic particles. American journal of respiratory and critical care medicine. 2009/01/08 ed. 2009 Apr 1;179(7):572–8.

49. Breton C V, Byun H-M, Wenten M, Pan F, Yang A, Gilliland FD. Prenatal Tobacco Smoke Exposure Affects Global and Gene-specific DNA Methylation. American Journal of Respiratory and Critical Care Medicine. 2009 Sep 1;180(5):462–7.

50. Schlinzig T, Johansson S, Gunnar A, Ekström TJ, Norman M. Epigenetic modulation at birth – altered DNA-methylation in white blood cells after Caesarean section. Acta Paediatrica. 2009 Jul 1;98(7):1096–9.

51. Newschaffer CJ, Croen LA, Fallin MD, Hertz-Picciotto I, Nguyen D V, Lee NL, et al. Infant siblings and the investigation of autism risk factors. Journal of Neurodevelopmental Disorders. 2012;4(1):7.

52. Hertz-Picciotto I, Schmidt RJ, Walker CK, Bennett DH, Oliver M, Shedd-Wise KM, et al. A Prospective Study of Environmental Exposures and Early Biomarkers in Autism Spectrum Disorder: Design, Protocols, and Preliminary Data from the MARBLES Study. Environ Health Perspect. 2018/11/23 ed. 2018;126(11):117004.

53. Bibikova M, Barnes B, Tsan C, Ho V, Klotzle B, Le JM, et al. High density DNA methylation array with single CpG site resolution. Genomics. 2011;98(4):288–95.

54. Aryee MJ, Jaffe AE, Corrada-Bravo H, Ladd-Acosta C, Feinberg AP, Hansen KD, et al. Minfi: A flexible and comprehensive Bioconductor package for the analysis of Infinium DNA methylation microarrays. Bioinformatics. 2014 May 15;30(10):1363–9.

55. Triche TJ, Weisenberger DJ, Van Den Berg D, Laird PW, Siegmund KD. Low-level processing of Illumina Infinium DNA Methylation BeadArrays. Nucleic Acids Research. 2013 Apr;41(7).

56. Chen Y, Lemire M, Choufani S, Butcher DT, Grafodatskaya D, Zanke BW, et al. Discovery of cross-reactive probes and polymorphic CpGs in the Illumina Infinium HumanMethylation450 microarray. Epigenetics. 2013 Feb;8(2):203–9.

57. Pidsley R, Zotenko E, Peters TJ, Lawrence MG, Risbridger GP, Molloy P, et al. Critical evaluation of the Illumina MethylationEPIC BeadChip microarray for whole-genome DNA methylation profiling. Genome Biology. 2016 Oct 7;17(1).

58. Gervin K, Salas LA, Bakulski KM, van Zelm MC, Koestler DC, Wiencke JK, et al. Systematic evaluation and validation of reference and library selection methods for deconvolution of cord blood DNA methylation data. Clin Epigenetics. 2019/08/27 ed. 2019;11(1):125.

59. Teschendorff AE, Breeze CE, Zheng SC, Beck S. A comparison of reference-based algorithms for correcting cell-type heterogeneity in Epigenome-Wide Association Studies. BMC Bioinformatics. 2017 Dec 13;18(1):105.

60. Yuan V, Hui D, Yin Y, Penaherrera MS, Beristain AG, Robinson WP. Cell-specific characterization of the placental methylome. BMC Genomics. 2021/01/06 ed. 2021;22(1):6.

61. Hansen KD. IlluminaHumanMethylation450kanno.ilmn12.hg19: Annotation for Illumina’s 450k methylation arrays. 2016.

62. Hansen KD. IlluminaHumanMethylationEPICanno.ilm10b2.hg19: Annotation for Illumina’s EPIC methylation arrays. 2016.

63. Abecasis GR, Auton A, Brooks LD, DePristo MA, Durbin RM, Handsaker RE, et al. An integrated map of genetic variation from 1,092 human genomes. Nature. 2012;491(7422):56–65.

64. Anderson CA, Pettersson FH, Clarke GM, Cardon LR, Morris AP, Zondervan KT. Data quality control in genetic case-control association studies. Nat Protoc. 2010/08/26 ed. 2010;5(9):1564–73.

65. Ritchie ME, Phipson B, Wu D, Hu Y, Law CW, Shi W, et al. limma powers differential expression analyses for RNA-sequencing and microarray studies. Nucleic acids research. 2015 Apr;43(7):e47.

66. Phipson B, Maksimovic J, Oshlack A. missMethyl: an R package for analyzing data from Illumina’s HumanMethylation450 platform. Bioinformatics. 2016 Sep 30;32(2):286–8.

67. Breeze CE, Reynolds AP, van Dongen J, Dunham I, Lazar J, Neph S, et al. eFORGE v2.0: updated analysis of cell type-specific signal in epigenomic data. Bioinformatics. 2019 Nov 1;35(22):4767–9.

68. Mordaunt CE, Jianu JM, Laufer BI, Zhu Y, Hwang H, Dunaway KW, et al. Cord blood DNA methylome in newborns later diagnosed with autism spectrum disorder reflects early dysregulation of neurodevelopmental and X-linked genes. Genome Med. 2020/10/14 ed. 2020;12(1):88.

69. Zhu Y, Mordaunt CE, Yasui DH, Marathe R, Coulson RL, Dunaway KW, et al. Placental DNA methylation levels at CYP2E1 and IRS2 are associated with child outcome in a prospective autism study. Hum Mol Genet. 2019;28(16):2659–74.

70. Zhu Y, Gomez JA, Laufer BI, Mordaunt CE, Mouat JS, Soto DC, et al. Placental methylome reveals a 22q13.33 brain regulatory gene locus associated with autism. Genome Biol. 2022 Feb 16;23(1):46.

71. Laufer BI, Hwang H, Jianu JM, Mordaunt CE, Korf IF, Hertz-Picciotto I, et al. Low-pass whole genome bisulfite sequencing of neonatal dried blood spots identifies a role for RUNX1 in Down syndrome DNA methylation profiles. Hum Mol Genet. 2021;29(21):3465– 76.

72. Krueger F, Andrews SR. Bismark: a flexible aligner and methylation caller for Bisulfite-Seq applications. Bioinformatics. 2011/04/14 ed. 2011;27(11):1571–2.

73. Coulson RL, Yasui DH, Dunaway KW, Laufer BI, Vogel Ciernia A, Zhu Y, et al. Snord116-dependent diurnal rhythm of DNA methylation in mouse cortex. Nat Commun. 2018/04/24 ed. 2018;9(1):1616.

74. Krueger F. Trim Galore!: A wrapper tool around Cutadapt and FastQC to consistently apply quality and adapter trimming to FastQ files. Babraham Inst.; 2015.

75. Langmead B, Salzberg SL. Fast gapped-read alignment with Bowtie 2. Nat Methods. 2012/03/04 ed. 2012;9(4):357–9.

76. Li H, Handsaker B, Wysoker A, Fennell T, Ruan J, Homer N, et al. The Sequence Alignment/Map format and SAMtools. Bioinformatics. 2009/06/08 ed. 2009;25(16):2078–9.

77. Ewels P, Magnusson M, Lundin S, Käller M. MultiQC: summarize analysis results for multiple tools and samples in a single report. Bioinformatics. 2016/06/16 ed. 2016;32(19):3047–8.

78. Korthauer K, Chakraborty S, Benjamini Y, Irizarry RA. Detection and accurate false discovery rate control of differentially methylated regions from whole genome bisulfite sequencing. Biostatistics. 2019;20(3):367–83.

79. Hansen KD, Langmead B, Irizarry RA. BSmooth: from whole genome bisulfite sequencing reads to differentially methylated regions. Genome Biol. 2012/10/03 ed. 2012;13(10):R83.

