## Supplemental Material for "Prenatal vitamin intake in first month of pregnancy and DNA methylation in cord blood and placenta in two prospective cohorts"

**Supplementary Material**

This supplementary material contains 5 supplementary figures and 1 supplementary table. An additional 6 large supplementary tables are provided as comma separated value tables.

**Supplemental Figure 1.** Sample exclusion/inclusion.

**MARBLES Cord Blood**

N=247

**MARBLES Placenta**

N=92

N=239

N=236

N=223

N=92

N=92

N=90

N=201

N=70

**EARLI**

**Cord Blood**

N=175

**EARLI Placenta**

N=134

Multiple Birth

N=2

N=173

N=170

N=170

N=128

N=127

N=127

N=113

N=88

Multiple Birth

N=8

Multiple Birth

N=6

Multiple Birth

N=0

Missing Variables of Interest

N=39

Missing Variables of Interest

N=57

Missing Variables of Interest

N=20

Missing Variables of Interest

N=22

Drop All But One Siblings

N=1

Drop All But One Siblings

N=0

Drop All But One Siblings

N=2

Drop All But One Siblings

N=13

Sex Mismatch

N=1

Sex Mismatch

N=3

Sex Mismatch

N=0

Sex Mismatch

N=3

**Supplemental Figure 2.** CpG probe exclusion/inclusion.

Fail Detection-P in >10% of Samples

N=661

Fail Detection-P in >5% of Samples

N=4,630

Fail Detection-P in >5% of Samples

N=1,699

Cross Reactive Probes

N=42,967

Cross Reactive Probes

N=43,068

Cross Reactive Probes

N=29,153

**MARBLES Cord Blood**

N=865,859

**MARBLES Placenta**

N=865,859

N=861,229

N=818,262

Remaining Y Chromosome Probes

N=379

N=817,883

N=864,160

N=821,092

N=821,008

Remaining Y Chromosome Probes

N=84

**EARLI Placenta and Cord**

N= 485,512

N= 484,851

N= 455,698

N= 455,650

Remaining Y Chromosome Probes

N=48

**Supplemental Figure 3.** Differences in array wide mean DNA methylation, split by regions in relation to CpG islands, comparing prenatal vitamin intake in month one of pregnancy versus no prenatal vitamin intake in month one of pregnancy. Regression models were adjusted for sex, maternal age, gestational age, maternal education, ancestry PCs, and estimated cell proportions.

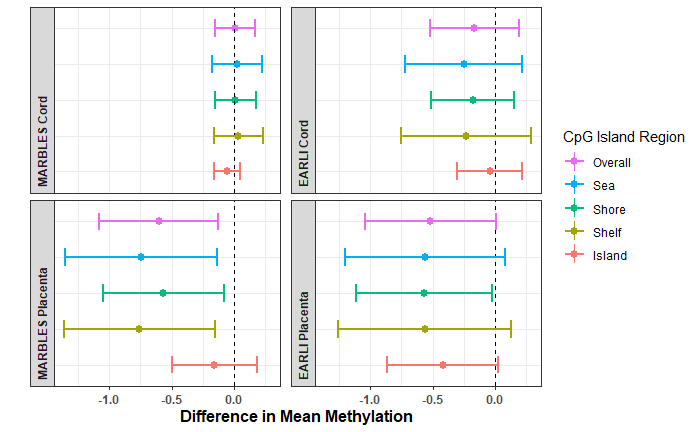

**Supplemental Figure 4.** Scatter plots of effect estimates for CpGs with P<0.01 in both tissues in **A)** MARBLES (n_CpGs_ =20) and **B)** EARLI (n_CpGs_ =66).

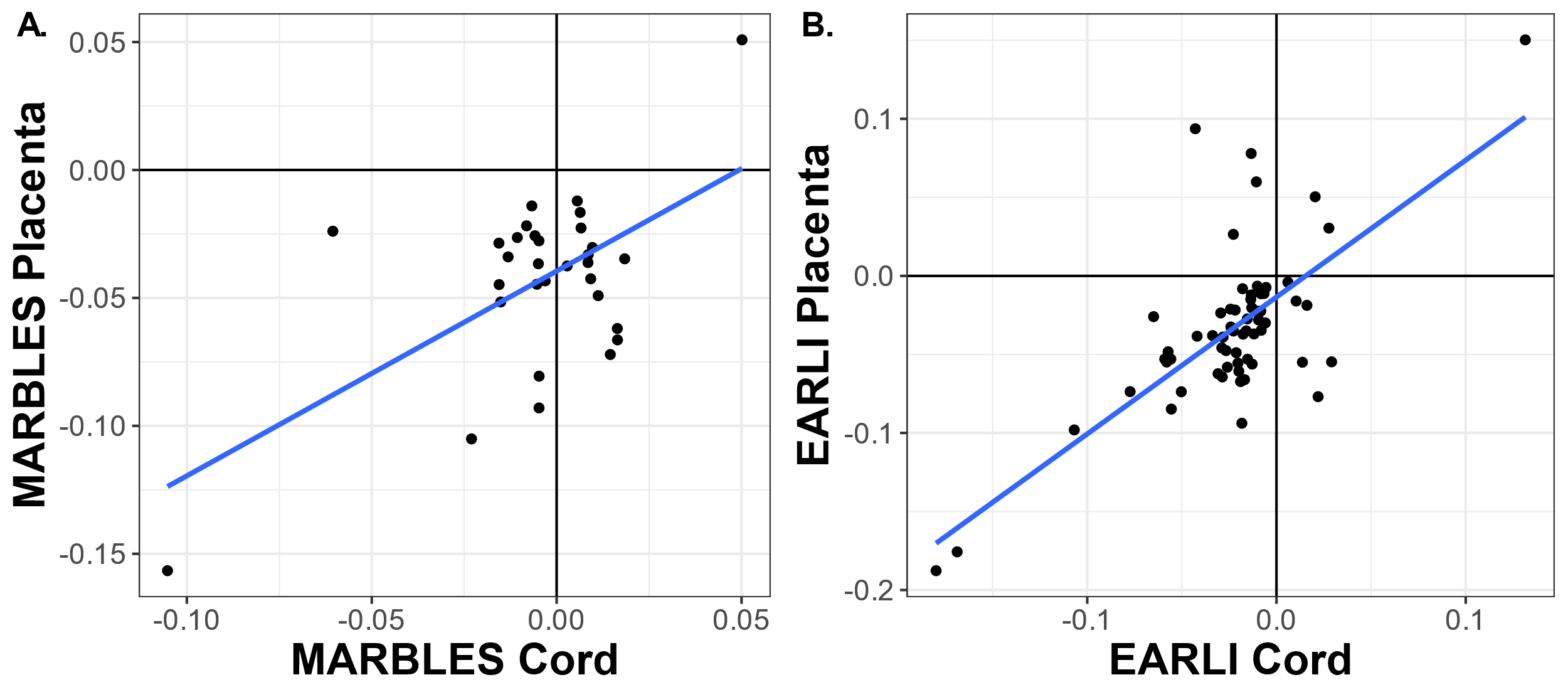

**Supplemental Figure 5.** eFORGE analysis for enrichment of chromatin state marker signatures. Dark points have adjusted-P value < 0.01, light points have adjusted-P > 0.01. (Abbreviations: ZNF/Rpts, zinc finger genes & repeats; TxWk, Weak transcription; TxFlnk, transcription at gene 5' and 3'; Tx, Strong transcription; TssBiv, Bivalent/Poised transcription start site; TssAFlnk, Flanking Active transcription start site; TssA, Active transcription start site; ReprPCWk, Weak Repressed PolyComb; ReprPC, Repressed PolyComb; Quies, Quiescent/Low; Het, Heterochromatin; EnhG, Generic Enhancers; EnhBiv, Flanking Bivalent TSS/Enhancer; Enh, Enhancers; BivFlnk, Flanking Bivalent TSS/Enh).

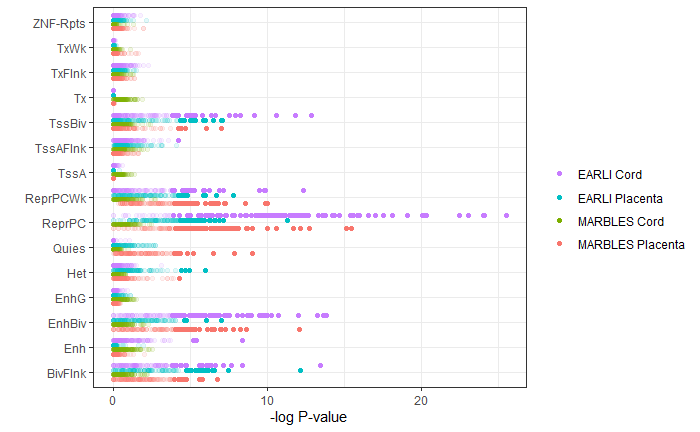

**Supplemental Table 7.** Whole-genome bisulfite sequencing (WGBS) differentially methylated region (DMR) analysis compared with methylation array results

|  | **MARBLES Placenta WGBS (n=39 with PV, 52 no PV)** | **MARBLES Cord Batch 1 WGBS (n=17 with PV, 28 no PV)** | **MARBLES Cord Batch 2 WGBS (n=17 with PV, 25 o PV)** | **EARLI Cord WGBS (33 with PV, 30 no PV)** |
| --- | --- | --- | --- | --- |
| **N DMRs in WGBS** | 165 | 301 | 469 | 36 |
| **Lower Methylation DMRs in WGBS** | 132 (80.0%) | 150 (49.8%) | 319 (68.0%) | 23 (63.9%) |
| **Array CpGs within DMRs in WGBS** | 61 | 65 | 120 | 4 |
| **Array CpGs within DMRs in same direction of effect as WGBS DMR** | 48 (78.7%) | 36 (55.4%) | 58 (48.3%) | 3 (75.0%) |
| **Array CpGs within 5kb of DMRs in WGBS** | 803 | 1243 | 2176 | 155 |
| **Array CpGs within 5kb of DMRs in same direction of effect as WGBS DMR** | 535 (66.6%) | 631 (50.8%) | 1061 (48.8%) | 91 (58.7%) |

**Other Supplemental Files:**

**Single site results spreadsheets for all CpGs**

**Supplemental Table 1.** Single site methylation array regression results for use of prenatal vitamins in the first month of pregnancy in placenta of EARLI cohort.

**Supplemental Table 2.** Single site methylation array regression results for sites use of prenatal vitamins in the first month of pregnancy in placenta of MARBLES cohort.

**Supplemental Table 3.** Single site methylation array regression results for use of prenatal vitamins in the first month of pregnancy in cord blood of EARLI cohort.

**Supplemental Table 4.** Single site methylation array regression results for use of prenatal vitamins in the first month of pregnancy in cord blood of MARBES cohort.

**Supplemental Table 5.** Gene ontology ranking by p-value for EARLI placenta, EARLI cord blood, MARBLES placenta, and MARBLES cord blood, ordered by rank sum across the four cohort/tissue groups.

**Supplemental Table 6.** Differentially methylation region analysis for whole genome bisulfite sequencing data
